# Short and Late Term Mortality and Quality of Life in Patients Requiring Continuous Veno-Venous Hemofiltration after Cardiac Surgery

**DOI:** 10.1101/2024.12.30.24319768

**Authors:** Minh-Thu Duong, Masood A Shariff, Fady G Haddad, Paul Catella, Chadi Salmane, Seleshi Demissie, Elie El-Charabaty, Suzanne El-Sayegh, Joseph T McGinn, John P Nabagiez

## Abstract

**Purpose:** Acute kidney injury (AKI) is a significant cause of morbidity and mortality post cardiac surgery sometimes requiring intermittent hemodialysis (IHD) or continuous veno-venous hemofiltration (CVVH). Previous studies included only procedures utilizing cardiopulmonary bypass (CPB). We reviewed CPB and non-CPB patients who developed AKI and required CVVH versus no CVVH.

**Materials and Methods:** 141 patients undergoing adult cardiac surgery developed AKI. 70 required CVVH (group1) and 71 did not (group2). We compared 30 day mortality, renal recovery, respiratory status and late mortality between both groups.

**Results:** 73% of patients in group1 required CVVH compared to 51% in group2 (p=0.007). Mortality was 38% versus 25% respectively at 1year (p=0.23) and 54% versus 17% respectively at 30 days (p<0.0001). Renal recovery was found in 47% versus 85%, freedom from ventilator support in 59% versus 83%, and discharge to long term nursing facility (LTNF) in 69% versus 44% respectively.

**Conclusion:** Mortality was significantly higher in CVVH group at 30 days but not at 1 year suggesting a long term benefit in those who survive the acute episode. The CVVH group demonstrated a higher rate of discharge to LTNF and lower rates of full renal recovery and freedom from ventilator support at discharge.

## Introduction

Acute kidney injury (AKI) is a significant cause of morbidity and mortality following cardiac surgery, reported in up to 30% of cardiac surgery patients depending on the definition used (1,2). The treatment for AKI following cardiac surgery includes judicious fluid management, intermittent hemodialysis (IHD) and continuous veno-venous hemofiltration (CVVH). CVVH is preferred over IHD to preserve stability of volume status (3) however CVVH is costly, resource intensive and may not improve outcome (4,5). Studies comparing the mortality of IHD and CVVH in cardiac surgery produce varying results regarding early and late outcomes. Thus far, studies have only included patients requiring cardiopulmonary bypass (CPB). Our study reviews CPB and non-CPB patients who developed AKI, comparing prognosis and quality of life at one month and one year.

We sought to assess rates of 30 day and one-year mortality, ventilator dependence, permanent hemodialysis and discharge to long-term nursing facility for CVVH and non-CVVH patients with AKI. We anticipated worse outcomes for the CVVH group based on the current literature and practice, where CVVH is utilized in patients with hemodynamic instability (6), and that type of surgery, urgency, the utilization of CPB, and postoperative complications before initiation of CVVH or IHD all greatly influence outcome.

## Methods

### Study Population and Design

This is a retrospective review of 2998 adults who underwent cardiac surgery between January 2005 and October 2013 at a single center. Patients with preoperative end stage renal disease on hemodialysis were excluded. Patients who developed postoperative AKI were included. AKI was defined as an increase in serum creatinine by ≥0.3 mg/dl within 48 hours, increase in serum creatinine to ≥1.5 times baseline, or urine volume <0.5ml/kg/h for 6 hours post cardiac surgery (7). Patients were classified into 2 groups: Group 1 required CVVH and Group 2 required IHD or no dialysis. Preoperative risk factors were measured by EuroSCORE II and Parsonnet mortality risk. The type of surgery, urgency of surgery and the utilization of CPB were noted. Postoperative complications prior to the initiation of CVVH or IHD were categorized as cardiac, pulmonary, neurological, infectious, vascular and gastrointestinal. Units of blood transfused in the first 7 days postoperatively and length of mechanical ventilation were also noted. Primary outcomes were 30 day mortality and late mortality defined as any death occurring in postoperative months two through twelve. Secondary outcomes for quality of life after hospital discharge included renal recovery, need for long-term mechanical ventilation, and disposition, including home, short-term rehabilitation or long-term nursing facility (LTNF).

### Statistical Analysis

Demographic and clinical baseline characteristics were summarized by study group. Categorical variables were summarized by frequencies and percentages and continuous variables were summarized using mean ± standard deviation (SD). The statistical analysis for comparison of categorical outcome variables between study groups (subjects who had CVVH and subjects who did not have CVVH) was performed using Chi-square test or Fisher’s exact test, as appropriate. Differences between study groups in continuous variables were analyzed with independent sample t test or nonparametric Mann-Whitney U test. Proportion of patient survival was assessed using the Kaplan–Meier method and compared with the log-rank test, at 30 days and past 30 days up to one year (deaths were censored in both). All analyses were two-sided and a P-value of <0.05 was considered to indicate statistical significance. All statistical analyses were performed using the SAS software, Version 9.3 (SAS Inc., Cary, NC, USA).

## Results

### Patient Characteristics

Of the 141 patients who developed AKI, 50% required CVVH (n=70). Baseline characteristics between the two groups are provided in Table 1. Regarding Groups 1 and 2: mean age was 71 ± 11 and 71 ± 10 years (p=0.804); males were 60% and 76% (p=0.048); chronic obstructive pulmonary disease 33% (n=23) and 24% (n=17) (p=0.240); and congestive heart failure 61% (n=43) and 49% (n=35) (p=0.147), respectively. There was no significant difference regarding preoperative stages of chronic kidney disease (CKD). Proteinuria in Groups 1 and 2 was 30% (n=21) and 46% (n=33), respectively (p=0.044).

**Table 1.**
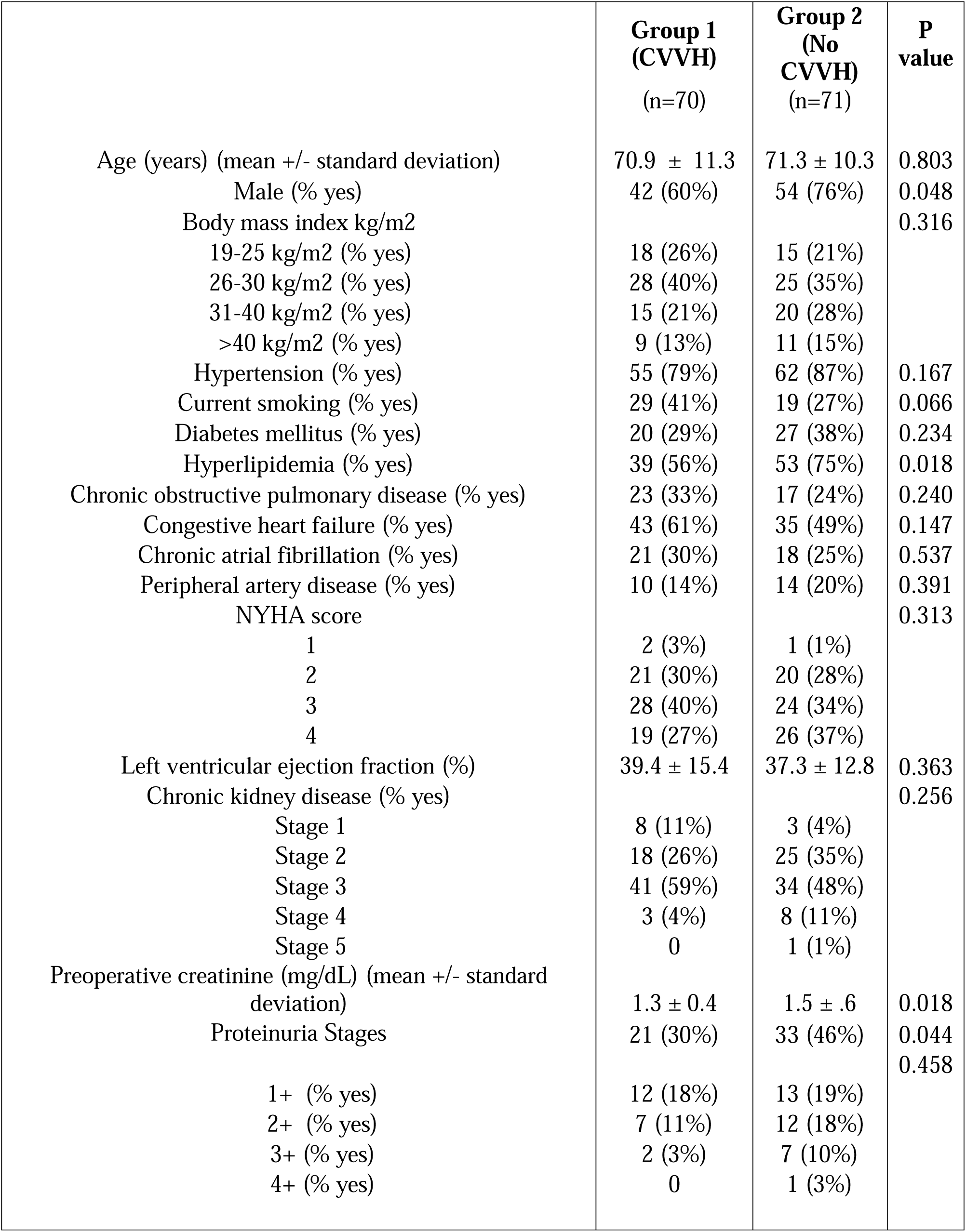
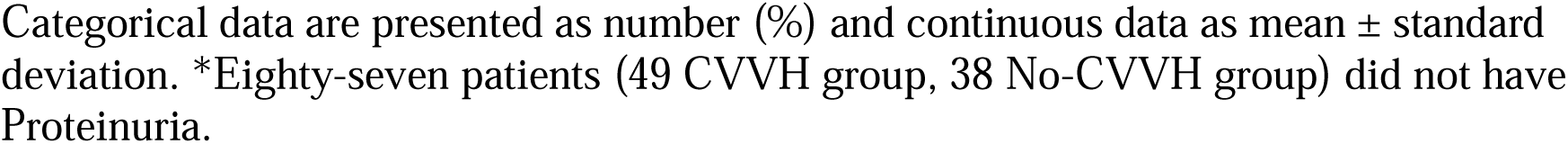
Baseline characteristics.

### Study Outcomes

Mean EuroSCORE II risk for Groups 1 and 2 was 30% ± 23 and 23% ± 19, respectively (p=0.061) and mean Parsonnet score risk was 26% ± 24 and 19% ± 16, respectively (p=0.113). Operative times for Groups 1 and 2 were 370 ± 135 and 328 ± 119 minutes (p=0.052), respectively; CPB was utilized in 73% (n=51) and 51% (n=36), for Groups 1 and 2, respectively (p=0.007). Isolated CABG was performed off pump in 64% of Group 1 (18/28) and 85% of Group 2 (34/40) compared to those utilizing CPB with beating heart (“pump assist”) in 36% for Group 1 (10/28) and 15% for Group 2 (6/40), p=0.080. Preoperative risk factors are detailed in Table 2.

**Table 2.**
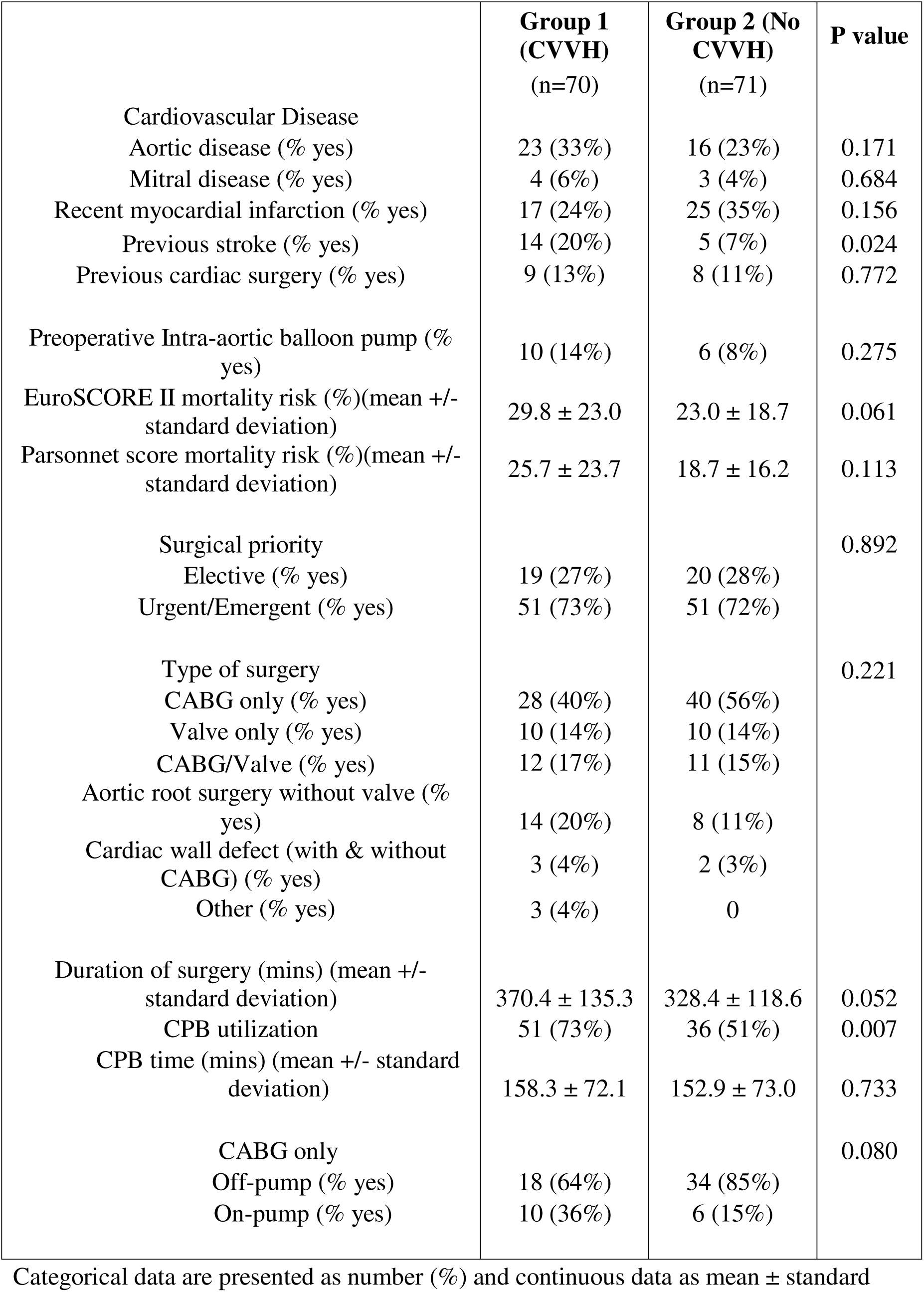
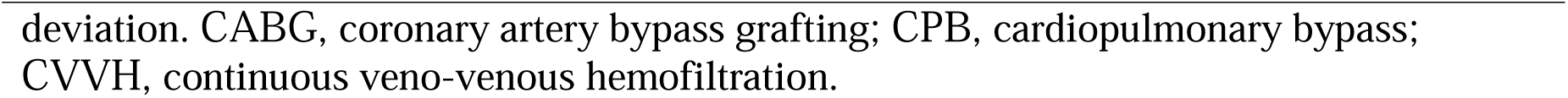
Preoperative and intraoperative risk factors.

|  | **Group 1**<br>**(CVVH)**<br>**(n=70)** | **Group 2 (No**<br>**CVVH)**<br>**(n=71)** | **P value** |
| --- | --- | --- | --- |
| Cardiovascular Disease |  |  |  |
| Aortic disease (% yes) | 23 (33%) | 16 (23%) | 0.171 |
| Mitral disease (% yes) | 4 (6%) | 3 (4%) | 0.684 |
| Recent myocardial infarction (% yes) | 17 (24%) | 25 (35%) | 0.156 |
| Previous stroke (% yes) | 14 (20%) | 5 (7%) | 0.024 |
| Previous cardiac surgery (% yes) | 9 (13%) | 8 (11%) | 0.772 |
| Preoperative Intra-aortic balloon pump (%<br>yes) | 10 (14%) | 6 (8%) | 0.275 |
| EuroSCORE II mortality risk (%)(mean +/-<br>standard deviation) | 29.8 ± 23.0 | 23.0 ± 18.7 | 0.061 |
| Parsonnet score mortality risk (%)(mean +/-<br>standard deviation) | 25.7 ± 23.7 | 18.7 ± 16.2 | 0.113 |
| Surgical priority |  |  | 0.892 |
| Elective (% yes) | 19 (27%) | 20 (28%) |  |
| Urgent/Emergent (% yes) | 51 (73%) | 51 (72%) |  |
| Type of surgery |  |  | 0.221 |
| CABG only (% yes) | 28 (40%) | 40 (56%) |  |
| Valve only (% yes) | 10 (14%) | 10 (14%) |  |
| CABG/Valve (% yes) | 12 (17%) | 11 (15%) |  |
| Aortic root surgery without valve (%<br>yes) | 14 (20%) | 8 (11%) |  |
| Cardiac wall defect (with & without<br>CABG) (% yes) | 3 (4%) | 2 (3%) |  |
| Other (% yes) | 3 (4%) | 0 |  |
| Duration of surgery (mins) (mean +/-<br>standard deviation) | 370.4 ± 135.3 | 328.4 ± 118.6 | 0.052 |
| CPB utilization | 51 (73%) | 36 (51%) | 0.007 |
| CPB time (mins) (mean +/- standard<br>deviation) | 158.3 ± 72.1 | 152.9 ± 73.0 | 0.733 |
| CABG only |  |  | 0.080 |
| Off-pump (% yes) | 18 (64%) | 34 (85%) |  |
| On-pump (% yes) | 10 (36%) | 6 (15%) |  |

Categorical data are presented as number (%) and continuous data as mean ± standard
deviation. CABG, coronary artery bypass grafting; CPB, cardiopulmonary bypass; CVVH, continuous veno-venous hemofiltration.

Statistically significant postoperative complications for Groups 1 and 2, before the initiation of CVVH or IHD, were: infectious 47% and 14% (p<0.001), cardiac 71% and 56% (p=0.062) and vascular 13% and 3% (p=0.026). Mean units of packed red blood cells (PRBC) transfused was 8.5±6.1 and 6.5±7.1 units (p=0.083) and mean ventilator days was 13±21 and 6±14 (p<0.0001) for Groups 1 and 2, respectively. Postoperative risk factors are detailed in Table 3. The average number of postoperative days to initiation of CVVH in Group 1 was 4.3±6.7 days and the mean duration of continuous renal replacement therapy was 4.3±3.9 days (Table 3).

**Table 3.** Postoperative risk factors and CVVH parameters.

|  | **Group 1**<br>**(CVVH)**<br>**(n=70)** | **Group 2**<br>**(No**<br>**CVVH)**<br>**(n=71)** | **P value** |
| --- | --- | --- | --- |
| Postoperative complications |  |  |  |
| Infectious (% yes) | 33 (47%) | 10 (14%) | 0.001 |
| Pulmonary (% yes) | 28 (40%) | 20 (28%) | 0.138 |
| Gastrointestinal (% yes) | 26 (37%) | 21 (30%) | 0.341 |
| Cardiac (% yes) | 50 (71%) | 40 (56%) | 0.062 |
| Neurologic (% yes) | 4 (6%) | 5 (7%) | 0.747 |
| Vascular (% yes) | 9 (13%) | 2 (3%) | 0.026 |
| Other (% yes) | 5 (7%) | 0 | 0.022 |
| Reoperation (% yes) | 15 (21%) | 13 (18%) | 0.643 |
| Transfused units of red blood cells (units) (mean<br>+/- standard deviation) | 8.5 ± 6.1 | 6.5 ± 7.1 | 0.083 |
| Days on mechanical ventilation (mean +/- standard<br>deviation) | 13.2 ± 21.3 | 5.9 ± 14.3 | 0.001 |
| Days to Initiate CVVH after development of AKI<br>(days) (Mean +/- standard deviation) | 4.3 ± 6.7 |  |  |
| Pre-CVVH Blood urea nitrogen level (mg/dL)<br>(mean +/- standard deviation) | 50.7 ± 31.2 |  |  |
| Post-CVVH Blood urea nitrogen level (mg/dL)<br>(mean +/- standard deviation) | 45.8 ± 24.3 |  |  |
| Pre-CVVH Creatinine (mg/dL) (mean +/- standard<br>deviation) | 3.3 ± 6.6 |  |  |
| Post-CVVH Creatinine (mg/dL) (mean +/-<br>standard deviation) | 2.6 ± 1.2 |  |  |
| Days on continuous renal replacement therapy<br>(days) (Mean +/- standard deviation) | 4.3 ± 3.9 |  |  |

Categorical data are presented as number (%) and continuous data as mean ± standard deviation. AKI, acute kidney injury; CVVH, continuous veno-venous hemofiltration.

The primary outcome for Groups 1 and 2 were: mean length of stay of 22±21 and 17±18 days (p=0.211) and 30 day mortality of 54% (38) and 17% (12) (p<0.0001), respectively. Primary and secondary outcomes are listed in table 4. In the patients who survived beyond 30 days, renal recovery was full for 45% (15) and 83% (50), temporary hemodialysis was 39% (13) and 5% (3) and permanent hemodialysis 15% (5) and 12% (7), respectively (p=<0.0001). Freedom from ventilator support at 30 days was 59% (19) and 82% (49), respectively (p=0.021). Survivors were discharged to: home 9% (3) and 39% (23), short term rehabilitation 24% (8) and 17% (10) and LTNF 67% (n=22) and 44% (n=26), respectively (p=0.006). For Group 1 and Group 2, mortality from postoperative months 2 through 12 was 38% (12/32) and 25% (15/59), respectively (p=0.229). Kaplan-Meier survival stratification was used for 30 day and late survival up to one year (Figure 1).

**Figure 1:**
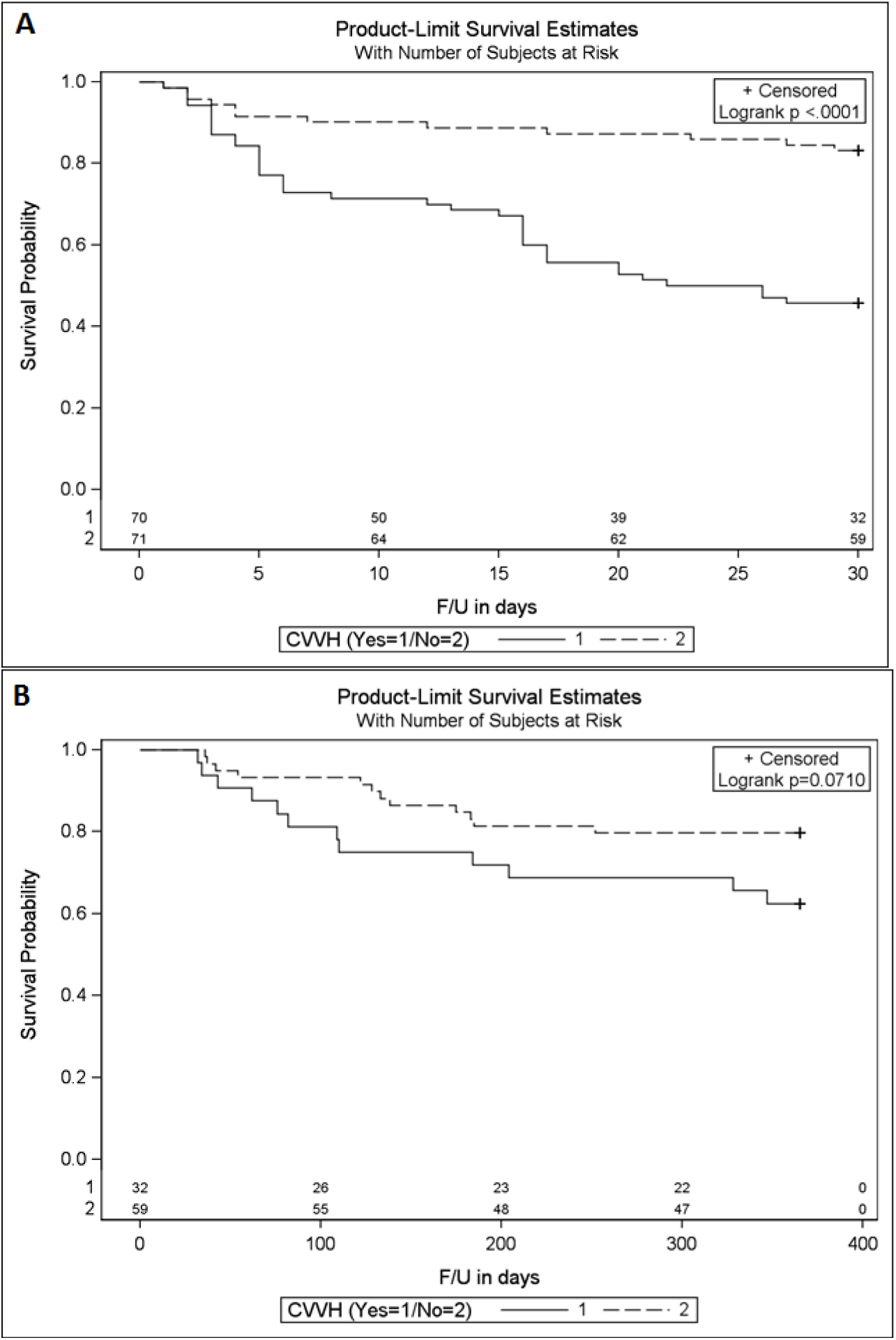
Kaplan-Meier survival by Group 1 (CVVH) and Group 2 (No CVVH) for postoperative 30 days (A) and late follow-up past 30 days up to one-year (B).

**Table 4.** Primary and secondary outcomes.

|  | **Group 1**<br>**(CVVH)**<br>**(n=70)** | **Group 2 (No**<br>**CVVH)**<br>**(n=71)** | **P value** |
| --- | --- | --- | --- |
| Hospital days (days) (mean +/- standard deviation) | 21.6 ± 21.0 | 16.7 ± 18.0 | 0.255 |
| 30 day mortality (% yes) | 38 (54%) | 12 (17%) | 0.001 |
| **Patients surviving 30 days (months 2 through 12 postop):** |  |  |  |
| Renal recovery |  |  | 0.001 |
| Full recovery (% yes) | 15 (45%) | 50 (83%) |  |
| Permanent hemodialysis (% yes) | 5 (15%) | 7 (12%) |  |
| Temporary hemodialysis (% yes) | 13 (39%) | 3 (5%) |  |
| Respiratory status |  |  | 0.021 |
| Spontaneous breathing (% yes) | 19 (59%) | 49 (82%) |  |
| Tracheostomy without ventilator assistance (% yes) | 13 (41%) | 11 (18%) |  |
| Disposition |  |  | 0.006 |
| Home (% yes) | 3 (9%) | 23 (39%) |  |
| Short term rehabilitation facility (% yes) | 8 (24%) | 10 (17%) |  |
| Nursing Home (% yes) | 22 (67%) | 26 (44%) |  |
| Mortality (months 2 through 12 postop) (% yes) | 12 (38%) | 15 (25%) | 0.229 |

Categorical data are presented as number (%) and continuous data as mean ± standard deviation. CVVH, continuous veno-venous hemofiltration.

## Discussion

Postoperative AKI in cardiac surgery is associated with a mortality of 40-90% (8). Even with standard IHD, the mortality rate with postoperative AKI remains high (9). During the study period, i.e. January 2005 and October 2013, 2998 adult patients who underwent cardiac surgery at Staten Island University Hospital, of whom 141 (4.7%) developed acute kidney injury. Compared to the acute renal kidney injury rate of 6.3% (16) and 6.7% (13) in previous studies, our reduced rate of AKI was proposed to be secondary to the inclusion of non-CPB patients. Our rate of post cardiac surgery patients requiring CVVH (2.3%) proved non inferior compared to 1.79% (15), 2.6% (17), 5% (13), 5% (16) in previous studies. This retrospective descriptive study adds to the existing literature demonstrating a difference in mortality and long-term prognosis between CVVH and IHD patients. Although it is impossible to assign causality, clearly these two modes of dialysis represent markers of the acuity of these patients.

The CVVH group had a statistically higher 30 day mortality. Mean lengths of stay and one-year mortality between CVVH and non-CVVH groups were not statistically significant. Mean length of stay in our CVVH group (22 days) was comparable to previous studies at 12.8 (17) and 24.5 (14) days. Quality of life in terms of renal recovery, freedom from ventilator dependence and disposition was significantly compromised in the CVVH group. Our CVVH group demonstrated similar rate of requiring permanent hemodialysis (15%) compared to a previous study at 15% (17). More patients in the CVVH group required permanent hemodialysis, tracheostomy and discharge to a long term nursing facility.

Preoperatively, patients in the CVVH group had a higher mean EuroScore II and Parsonnet score but the difference did not reach statistical significance. The CVVH group had a significantly higher rate of infectious, cardiac and vascular complications prior to initiation of hemodialysis. The CVVH group also had a higher mean number of PRBC units transfused within the first 7 days postoperatively and a significantly higher mean number of days on mechanical ventilation. These findings suggest that the CVVH group was more critically ill prior to initiation of hemofiltration.

The CVVH group also had a higher utilization of CPB which is associated with a systemic inflammatory response secondary to transient endotoxemia (10). On-pump procedures are linked to increased production of oxygen-derived free radicals, activation of neutrophils and complement cascade, all of which may negatively affect the heart, lung and kidneys (11). Regardless, hemodynamic instability requiring CPB for isolated CABG patients (beating heart “pump assist”) (12) predisposed to AKI requiring CVVH, unlike the off-pump CABG patients, who by definition would have maintained hemodynamic stability.

Mortality is known to be high in those requiring CVVH for AKI following cardiac surgery. Previous studies, all utilizing CPB, reported 30-day mortality rates at 35% (17), 38.8% (13), 40% (14), 42% (18), 43% (16), 55.5% (15). Our results are comparable, suggesting that the off pump component of our series did not appear to translate into significantly improved 30-day outcomes specifically in those requiring CVVH. Interestingly, although we found a difference in 30 day survival there was no statistically significant difference in survival for months 2 through 12 postoperatively (p=0.229), suggesting a benefit for CVVH in patients who survive beyond 30 days.

## Limitations

There are several limitations to this study. It is a retrospective review of a single center experience with a relatively small sample size, which limits generalizability. No definitive conclusions can be made regarding isolated CABG outcomes based on off-pump versus CPB utilization due to the selection bias introduced by intraoperative hemodynamics. Initiation of CVVH was not protocol driven, but rather chosen on a case-by-case analysis by a multi-disciplinary team of surgeons, intensivists and nephrologists. Because the study includes patients treated over a period of 8 years, specific practice patterns may have changed during the time period.

## Conclusion

This is the first study to include non-CPB patients and the largest study to assess late mortality and quality of life in CPB and non-CPB patients who developed AKI. Although 30 day mortality was significantly higher in the CVVH group, no difference was seen in one year mortality for those who survived the first month, suggesting a long term benefit in those who survive the acute episode. The CVVH group demonstrated lower rates of full renal recovery and freedom from ventilator support at discharge, as well as a higher rate of discharge to long-term nursing facility during the 30-day postoperative period.

## Data Availability

All data produced in the present work are contained in the manuscript

## Acknowledgements

We thank the volunteers who assisted with data collection Carmen Chan and Umair Khan.

## References

[1] Rosner MH, Okusa MD. Acute kidney injury associated with cardiac surgery. Clin J Am Soc Nephrol 2006;1(1):19–32.

[2] Karkouti K, Wijeysundera DN, Yau TM, Callum JL, Cheng DC, Crowther M, et al. Acute kidney injury after cardiac surgery: focus on modifiable risk factors. Circulation 2009;119:495–502.

[3] Bellomo R, Ronco C. Continuous versus intermittent renal replacement therapy in the intensive care unit. Kidney Int Suppl 1998;66:S125–8.

[4] Teehan GS, Liangos O, Jaber BL. Update on dialytic management of acute renal failure. J Intensive Care Med 2003;18:130–8.

[5] Tsang GM, Khan I, Dar M, Clayton D, Waller D, Patel RL. Hemofiltration in a cardiac intensive care unit: time for a rational approach. Asaio J 1996;42:M710–3.

[6] Hoste EA, Dhondt A. Clinical review: use of renal replacement therapies in special groups of ICU patients. Crit Care 2012;16:201.

[7] Pickering JW, Endre ZH. The definition and detection of acute kidney injury. J Renal Inj Prev 2013;3(1):21–5.

[8] Suen WS, Mok CK, Chiu SW, Cheung KL, Lee WT, Cheung D, et al. Risk factors for development of acute renal failure (ARF) requiring dialysis in patients undergoing cardiac surgery. Angiology 1998;49:789–800.

[9] Lange HW, Aeppli DM, Brown DC. Survival of patients with acute renal failure requiring dialysis after open heart surgery: early prognostic indicators. Am Heart J 1987;113:1138–43.

[10] Jansen NJ, van Oeveren W, Gu YJ, van Vliet MH, Eijsman L, Wildevuur CR. Endotoxin release and tumor necrosis factor formation during cardiopulmonary bypass. Ann Thorac Surg 1992;54:744–7; discussion 7-8.

[11] Laffey JG, Boylan JF, Cheng DC. The systemic inflammatory response to cardiac surgery: implications for the anesthesiologist. Anesthesiology 2002;97:215–52.

[12] Glauber M, Farneti A, Bevilacqua S, Karimov J. Pump-assisted beating heart surgery. Multimed Man Cardiothorac Surg 2007;2007:mmcts.2004.000943.

[13] Bapat V, Sabetai M, Roxburgh J, Young C, Venn G. Early and intensive continuous veno-venous hemofiltration for acute renal failure after cardiac surgery. Interact Cardiovasc Thorac Surg 2004;3:426–30.

[14] Bent P, Tan HK, Bellomo R, Buckmaster J, Doolan L, Hart G, et al. Early and intensive continuous hemofiltration for severe renal failure after cardiac surgery. Ann Thorac Surg 2001;71:832–7.

[15] Demirkiliç U, Kuralay E, Yenicesu M, Cağlar K, Oz BS, Cingöz F, et al. Timing of replacement therapy for acute renal failure after cardiac surgery. Journal of cardiac surgery 2004;19:17–20.

[16] Elahi MM, Lim MY, Joseph RN, Dhannapuneni RRV, Spyt TJ. Early hemofiltration improves survival in post-cardiotomy patients with acute renal failure. European J Cardiothorac Surg 2004;26:1027–31.

[17] Guclu O, Yavuz C, Gurkan SC, Yuksel V, Demirtas S, Caliskan A, et al. Continuous renal replacement therapy after cardiac surgery in patients with acute renal failure. Med Glas (Zenica) 2013;10:244–8.

[18] Vidal S, Richebé P, Barandon L, Calderon J, Tafer N, Pouquet O, et al. Evaluation of continuous veno-venous hemofiltration for the treatment of cardiogenic shock in conjunction with acute renal failure after cardiac surgery. Eur J Cardiothorac Surg 2009;36:572–9.

